# Importance and assessment of quality of life in symptomatic permanent atrial fibrillation: Patient focus groups from the RATE-AF trial

**DOI:** 10.1101/2020.08.21.20179077

**Authors:** Jacqueline Jones, Mary Stanbury, Sandra Haynes, Karina V Bunting, Trudie Lobban, A John Camm, Melanie J. Calvert, Dipak Kotecha, on behalf of the RAte control Therapy Evaluation in permanent Atrial Fibrillation (RATE-AF) trial group

**Author notes:** Address for correspondence: Dipak Kotecha MBChB MRCP PhD MSc FESC FHEA Professor of Cardiology, University of Birmingham Institute of Cardiovascular Sciences, Medical School, Vincent Drive, Birmingham, B15 2TT, UK. Joint first authors (Jacqueline Jones, Mary Stanbury).

## Abstract

**Aims:** To establish the extent and impact of symptoms in patients with atrial fibrillation (AF), the importance of different aspects of quality of life (QoL), and how we should assess wellbeing.

**Methods:** Focus groups of patients with symptomatic permanent AF in a trial of heart rate control; the RATE-AF trial randomised 160 patients aged ≥60 years with permanent AF and at least NYHA class II dyspnoea to either digoxin or beta-blockers. Patient and public representatives led the focus groups and performed all data acquisition and analysis, using thematic approaches to interpret patient views about QoL and its measurement.

**Results:** Substantial impairment of health-related QoL was noted in 160 trial patients, with impact on all domains apart from mental health. Eight women and 11 men aged 61–87 years participated in the focus groups. Common themes were a lack of information from healthcare professionals about AF, a lack of focus on QoL in consultations, and a sense of frustration, isolation and reduced confidence. There was marked variability in symptoms in individual patients, with some describing severe impact on activities of daily living, and profound interaction with comorbidities such as arthritis. Day-to-day variation in QoL and difficulty in attributing symptom burden to AF or other comorbidities led to challenges in questionnaire completion. Consensus was reached that collecting both general and AF-specific QoL would be useful in routine practice, along with participation in peer support, which was empowering for the patients.

**Conclusions:** The impact of comorbidities is poorly appreciated in the context of AF, with considerable variability in QoL that requires both generic and AF-specific assessment. Improvement in QoL should direct the appraisal, and reappraisal, of treatment decisions for patients with permanent AF.

## Introduction

Atrial fibrillation (AF) is a major burden on patients and healthcare services. These effects are projected to increase exponentially as our communities grow older and the incidence of AF in older people increases.^1^ Although adverse outcomes in AF such as stroke rightly receive attention from clinicians due to their preventable nature, poor patient quality of life (QoL) often lacks consideration in clinical practice, despite also being amenable to treatment.

Patients with AF have significantly poorer health-related QoL^2^, which includes comparison with both healthy individuals and those with other cardiovascular disease.^3^ This has been attributed to the variety of symptoms that AF patients can suffer, including lethargy, palpitations, dyspnoea, sleeping difficulties, chest discomfort and psychosocial distress, as well as anxiety related to treatments and potential complications.^4^ Although QoL is significantly related to mortality, AF-related symptoms do not necessarily track with clinical outcomes such as stroke, heart failure or myocardial infarction, making extraction of QoL information important for routine clinical management.^5^ The majority of published studies on QoL in AF relate to the response to rhythm control therapies such as antiarrhythmic drugs or ablation. In contrast, patients with permanent AF have even worse QoL^6^, account for around 50% of patients, and yet we lack adequate description of underlying factors.^7^

The current unknowns about QoL in patients with permanent AF limit the scope of how effective clinicians can be in addressing patient concerns. We designed a qualitative study, led by a Patient and Public Involvement (PPI) team, and embedded within a clinical trial; the RAte control Therapy Evaluation in permanent Atrial Fibrillation (RATE-AF) trial.^8^ Our aim was to explore three broad domains: (1) The perspective of patients on core components of health-related QoL, and how this is influenced by AF; (2) The measurement of QoL in AF and what tools were felt useful by patients to measure the response to treatment; and (3) Whether QoL was an important outcome that clinicians should address.

## Methods

This mixed methods study was part of the RATE-AF trial programme, a prospective, open-label, blinded end-point, randomised controlled trial of 160 patients with symptomatic permanent AF. The trial is the first to directly compare longer-term heart rate control using digoxin and beta-blocker therapy in this patient group. The rationale and design of the study have previously been described^7^; in brief, the trial was embedded within the UK National Health Service (NHS), with minimal selection criteria to reflect routine clinical care. Patients were aged 60 years or older with permanent AF in need of rate-control and breathlessness equivalent to at least New York Heart Association (NYHA) class II. As per guidelines, permanent AF was characterised as a physician decision for rate control with no plans for cardioversion, antiarrhythmic drugs or ablation.^9^ We only excluded patients with either clear requirements or contraindications for either drug, for example myocardial infarction in the last 6 months, history of severe bronchospasm, bradycardia or previous intolerance (http://clinicaltrials.gov NCT02391337, ISRCTN 95259705 and EudraCT 2015–005043–13). The RATE-AF trial and the qualitative aspects were sponsored by the University of Birmingham and funded by the National Institute for Health Research (NIHR).

### Patient and Public Involvement

A team of three PPI members helped to design and manage the trial, including positions on the Trial Steering Committee. The design of the focus groups was led by JJ and MS from the PPI team, with the support of cardiology, patient-reported outcomes research and qualitative research teams at the University of Birmingham.

### Participant recruitment and data collection

Patients for the RATE-AF trial were recruited from referrals to three hospital sites in Birmingham (Queen Elizabeth Hospital, City Hospital and Heartlands Hospital), and also directly from General Practices across the West Midlands region in the UK. As part of the consent procedures, all participants were asked if they could be contacted to contribute to the focus groups. From this cohort, 20 patients were consecutively invited to attend based on completion of all drug uptitration at that time (i.e. beyond the first 2 months of their trial participation), purposely sampled by gender and randomised group. No clinical variables were used to decide on focus group composition, but participants had to be able to attend on the specified date (five patients refused/were unable to attend on the date and were replaced with the next available patient). One patient who accepted was unable to attend on the day due to illness. Focus groups were held at the NIHR/Wellcome Trust Clinical Research Facility at the Queen Elizabeth Hospital Birmingham in 2019, and split into two meetings for each arm of the trial. The first meeting focused on building rapport within the group and the impact of AF on their lives, with the second meeting focused on assessment and tools to measure that impact. Each meeting lasted approximately 2.5 hours separated by two weeks (maximum of ten participants in each meeting). Focus groups were led by the patient and public representatives (JJ and MS), with DK also in attendance to address any medical issues.

### Reimbursement

Patients received a fixed sum of £50 for contribution to each focus group, in addition to appropriate compensation for travel and subsistence costs. PPI members received funding according to NIHR INVOLVE guidance (https://www.invo.org.uk/). There was no industry funding for any part of this trial.

### QoL tools

Three validated QoL questionnaires were used in the RATE-AF trial.^7^ AF-specific QoL was assessed using the Atrial Fibrillation Effect on QualiTy-of-life (AFEQT) questionnaire.^10^ Generic QoL was assessed using both the EuroQol EQ-5D-5L questionnaire^11^ and the Short Form 36 Health Survey (SF36).^12^ The SF36 survey is comprised of eight domains; normalised UK values were taken from 8,889 respondents of a large-scale social survey, the Third Oxford Health and Lifestyles Survey (OHLS-III), sampled from primary care in the UK.^13^ T-tests adjusted for unequal variance were used to compare mean values with data from the baseline visit of the RATE-AF trial. All participants of the focus groups had experience of completing all three questionnaires on at least 2 occasions during their trial visits. At the end of the first meeting, copies of the questionnaires were also provided to the participants for discussion at the next meeting.

### Data analysis

A topic guide was developed and finalised by this group prior to the focus groups, and used as a roadmap for discussions in each meeting. The topic guide included specific questions relating to the three study domains, and corresponding probe questions to explore these issues in more detail (**Supplementary file**). Audio recordings of the meetings were made with the consent of all participants. A professional service transcribed interviews (clean verbatim) and the transcription was reviewed for consistency and accuracy by JJ and MS. The data (recordings and transcripts) were analysed using thematic approaches.^14^ To organise the patient comments into the three key domains of interest, we developed a set of codes after the authors had familiarised themselves with the content of the transcripts. For the components of health-related QoL in AF, the codes related to any comment on: *Physical impact from AF; Emotional impact from AF; Daily life impact from AF; Impact on carers; Interaction with comorbidities; Prior knowledge of AF or lack thereof*; and *Importance of QoL to the patient*. For measurement of QoL in AF: *Value of the QoL questionnaire;* and *Comparison of QoL questionnaires*. For the importance of addressing QoL: *Treatment expectations; Impact of the trial medications on physical function;* and *Impact of the trial medications on QoL*. An iterative process was performed of coding each transcript (JJ and MS). The codes were compiled in a data spreadsheet with extracted statements given primary codes (main issue raised by the patient in that comment) and secondary codes (where, if required, an additional issue was raised in the same comment). JJ and MS also manually extracted specific quotes from patients that were relevant or particularly significant to the group as a whole, or may not have fit into the specified codes.

## Results

RATE-AF trial data at baseline from 160 patients with symptomatic permanent AF confirmed substantial reduction in QoL in all domains of the SF36 questionnaire, except for mental health. This was observed compared to the UK norm, as well as compared to those with a longstanding illness in population survey data (**Figure 1**).

**Figure 1:**
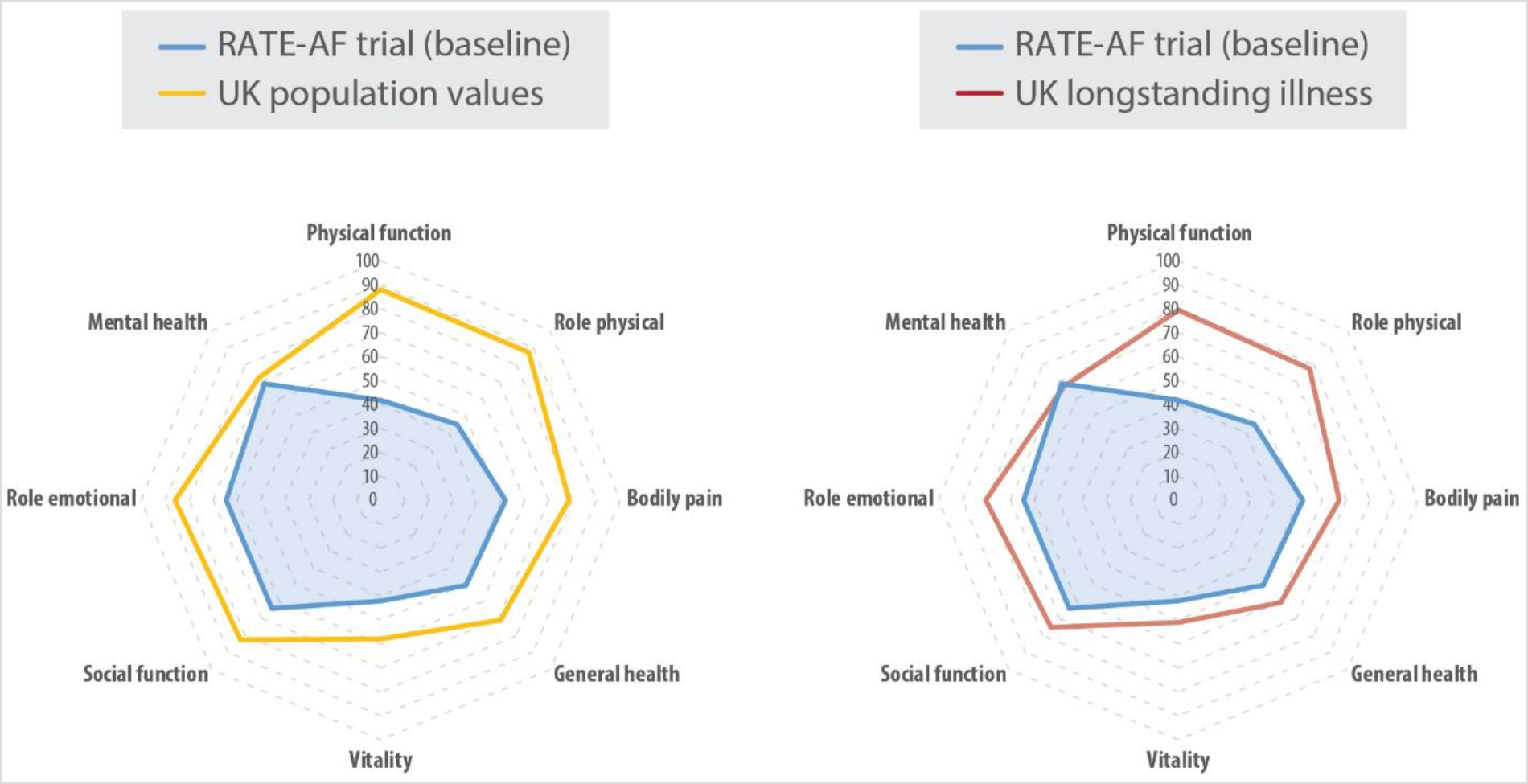
Comparative quality of life scores. SF36 quality of life scores in 160 RATE-AF patients. Left: Values compared to the UK population average from the OHLS-III social survey in 8,889 participants. Right: Comparison with the 40% of respondents declaring a longstanding illness in OHLS-III. Higher scores indicate better quality of life; all components demonstrated statistically worse quality of life in RATE-AF patients (p< 0.0001), except for mental health (p = 0.49).

Nineteen participants took part in the focus groups, comprising 8 women and 11 men all from a white British background. Mean age was 74 years (SD 7), with a range of 62–86 years. Patients had been diagnosed with AF for a mean of 5 years, and 10 (53%) had a known diagnosis of coexisting heart failure or signs of heart failure at baseline. Other common comorbidities were hypertension in 11 patients (58%), type 2 diabetes in 5 (26%), airways disease in 4 (21%), and previous stroke or transient ischaemic attack in 3 (16%). The demographics of the focus group participants were all comparable with the main trial population.^8^ All patients had completed uptitration of their trial medication (10 randomised to digoxin and 9 to beta-blockers), and were on stable dosage with adequate control of heart rate. A summary of key findings, determined by the Patient and Public leads, is presented in **Figure 2** and a summary for patients in **Figure 3**.

**Figure 2:**
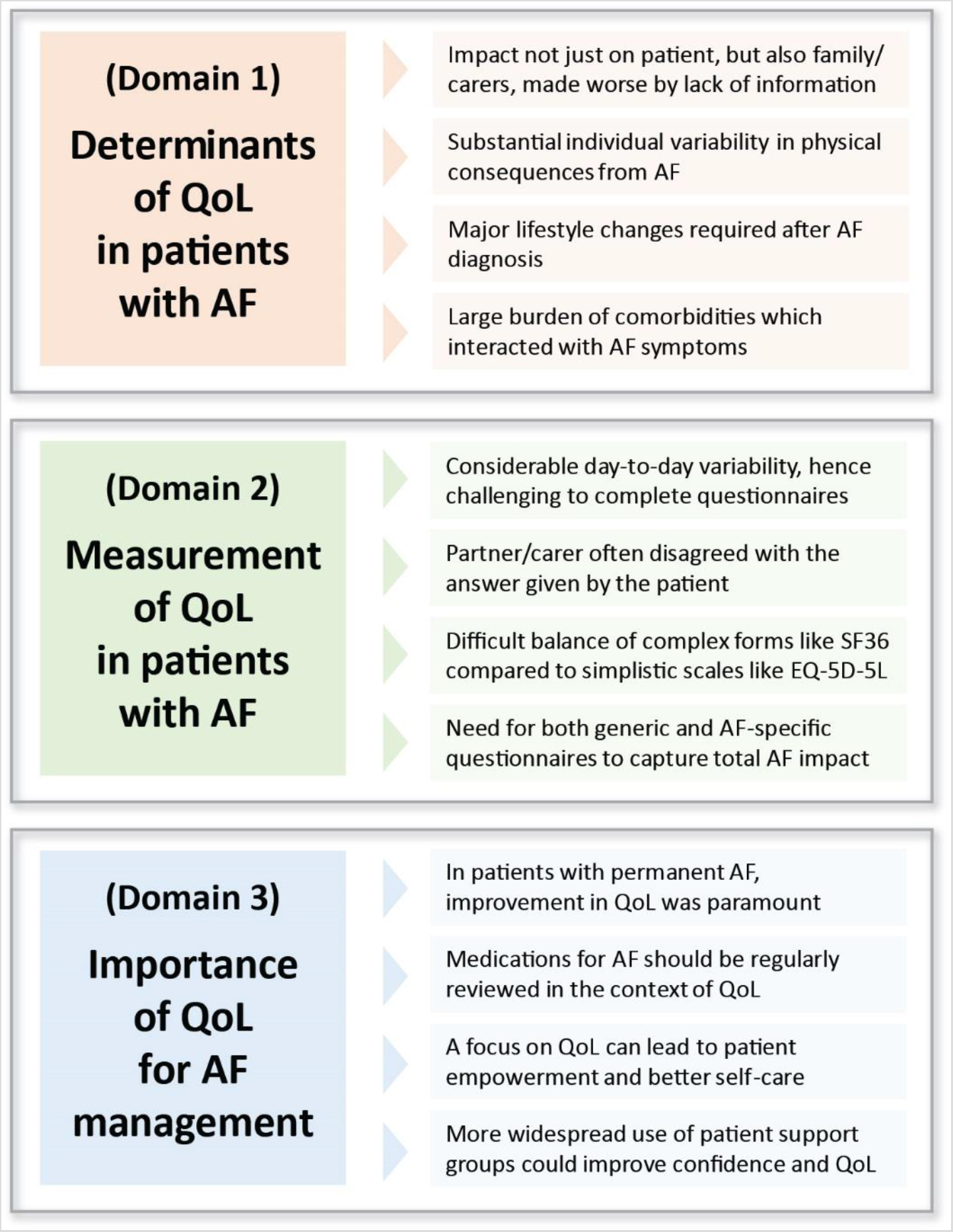
Summary of RATE-AF focus groups. Key issues raised by patients according to relevant themes. AF = Atrial fibrillation; QoL = Quality of life.

**Figure 3:**
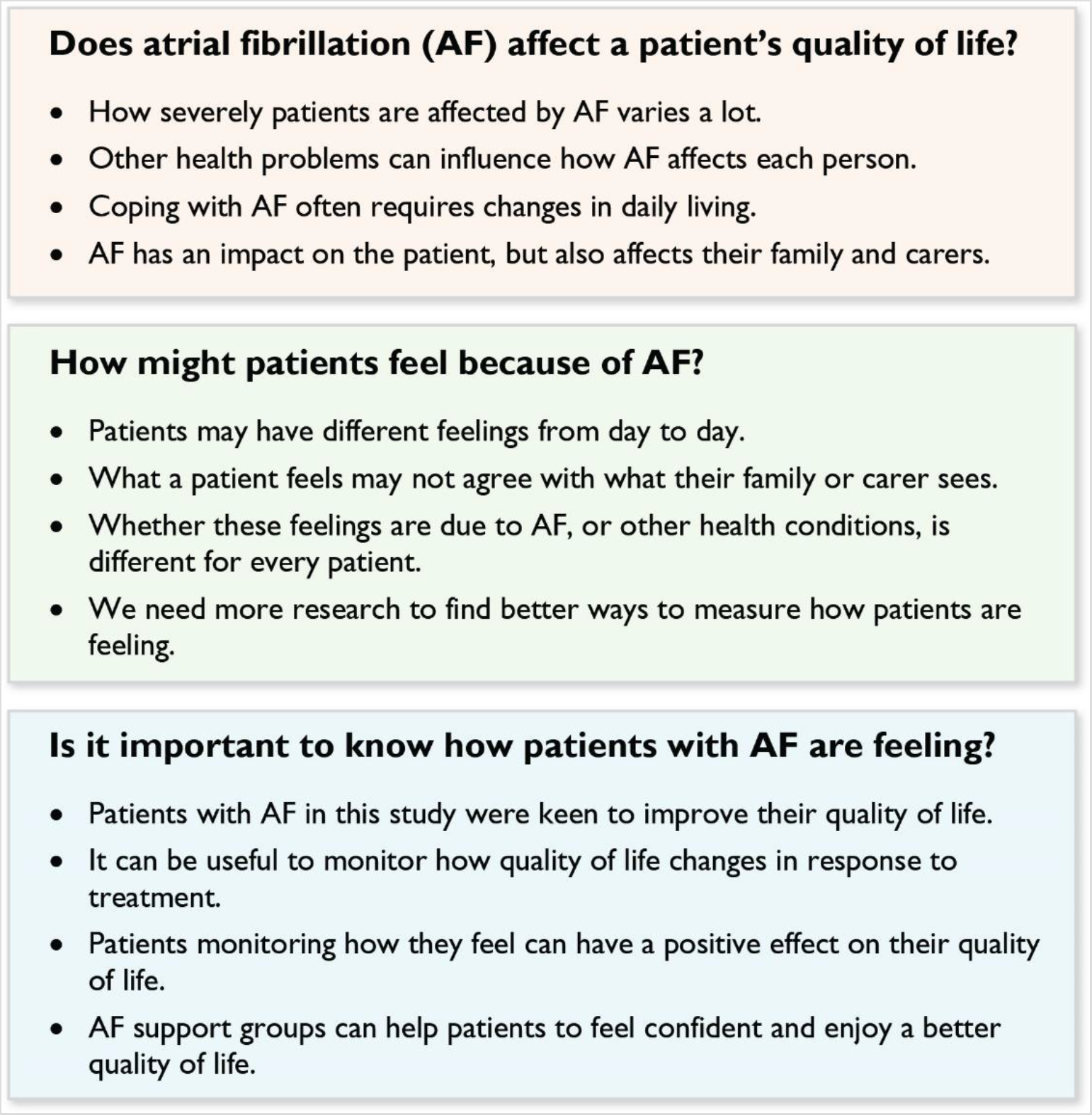
Summary for patients. Information about atrial fibrillation (AF) for patients can be found at: https://www.bhf.org.uk/informationsupport/conditions/atrial-fibrillation. Patient resources and support are available at: https://www.heartrhythmalliance.org/afa/uk/for-patients.

### (Domain 1) Determinants of QoL in patients with AF

Key issues raised with regards to determinants of QoL and how AF affects the lives of patients were:

1. There was consistent feedback about a lack of information from healthcare professionals about AF, and a lack of focus on QoL in healthcare consultations. This led to a sense of frustration, isolation and reduced confidence which contributed to the overall impact of AF on patient wellbeing.
2. The impact of AF was not just felt by patients, but also their primary caregivers and the wider family.
3. There was marked variability in AF symptoms in individual patients, with some describing severe impact on physical capacity and activities of daily living due to breathlessness, fatigue and dizziness, whereas others had few symptoms.
4. In group discussion, the patients concluded that the impact of comorbidities (especially large-joint arthritis) was the most important determinant of the effect of AF on physical activity. There was consistent feedback that the effect and treatment of comorbidities was poorly considered in interactions with healthcare professionals.
5. Adaptation to a new style of living was required after the diagnosis and treatment of AF, and there were considerable emotional impacts.

A summary of quotes from patients relevant to this domain are presented in **Table 1**.

**Table 1:** Determinants of QoL in patients with AF.

| Impact | Quote | Comments/context |
| --- | --- | --- |
| Emotional impact on patient and family | “As you say it can be a lonely thing [having AF]. You’ve got company round you but even so you are thinking am I the only one like this?”<br>“I’m losing my confidence to go out.”<br>“If you aren’t positive you might as well pack up and sit in your armchair.”<br>“Unfortunately my partner takes the brunt in the way I sometimes feel [anger and frustration]”<br>“I find the most exasperating thing is having to ask for help; I loathe it.”<br>“I sometimes think I don’t go anywhere where I’m more than half an hour away from [the hospital], because it’s frightening if something does go wrong.” | Common feelings were fear and bewilderment at the time of diagnosis, which persisted.<br>Consistent theme of inability or reticence to ask questions leading to a loss of confidence; often not verbalised to their usual healthcare practitioners.<br>General feelings of isolation, with a sense of loneliness that was pervasive despite interaction with family and friends.<br>Dependency and burden on caregivers was highlighted many times.<br>Fear of complications and the unknown was common, and impacted on overall QoL. |
| Impact on physical activity | “If I’m out on a walk and come to a hill it gets harder and harder and I get slower and slower.”<br>“I used to play rugby and cricket, but walking round the snooker table [is all I can manage] and I don’t know whether that counts [as exercise].”<br>“I feel a bit of a fraud because I have no symptoms. I walk 2.5 miles every day to get my paper, and I don’t just walk I pound it.”<br>“Housework and stuff - I can’t do what I used to do. When vacuuming sometimes I stop after five minutes and I’m breathless and [need to] sit down.”<br>“I can do my housework slowly and everything seems fine.”<br>“My life is so changed following [AF therapy] - I can walk and I’m a garden fanatic. I’m out there digging and sawing trees.” | There was a diverse range of comments on the impact of AF on physical capabilities.<br>The majority of patients felt that their AF had negatively impacted on physical activity, but a few patients had no apparent change in their ability. A minority recognised an increase in their ability to carry out physical activity due to medical therapy for previously undiagnosed AF.<br>The uncertainty of not knowing how they would feel on any given day was frustrating, particularly as causal factors were difficult to identify (i.e. whether due to AF, comorbidities, the general aging process or a combination).<br>The impact of emotional factors on physical activity was evident, particularly anxiety around their AF diagnosis, and the effect on their cardiovascular and general health. |
| Impact on daily living and wellbeing | “I feel degraded my wife has to shower me.”<br>“I was full of life, but no longer”.<br>“[I am] breathless and have no energy to stand there and do it [ironing]”.<br>“Previous lifestyle has now completely changed”.<br>“I force myself to do things. You can’t just sit in the chair all day looking out of the window”. | For a few patients, there was little change in daily living but the majority felt that their AF diagnosis had a major effect on daily well-being.<br>Living with the variability of symptoms on a daily basis created uncertainty for patients around being able to plan activities. There was a sense that on any given day their body would dictate the amount of activity they were able to undertake.<br>Asked to rate the impact of AF on their daily life (where 1 was no impact and 10 the greatest impact), the majority gave a score of 7 or above.<br>Some patients acknowledged that their previous lifestyle was not compatible with their current status after the diagnosis of AF. The realisation that significant changes were required in their daily planning was found to be extremely hard to accept and to adapt. |

### (Domain 2) Measurement of QoL in patients with AF

The key points discussed about tools to measure QoL and treatment response were:

1. Most patients found it difficult to complete questionnaires due to considerable day-to-day variation in their AF-related symptoms. There were challenges for questionnaires like EQ-5D-5L (which asks about impact today), as well as SF36/AFEQT (4-week recall).
2. Separation of emotional from physical well-being (for example in SF36) was confusing for many patients who felt that AF impacts were inter-related.
3. AFEQT had the most relevance to AF symptom burden, but attribution to AF or other comorbidities was challenging.
4. There was a difficult balance to strike between content and time taken; for example comparing the brevity of the EQ-5D-5L questionnaire with the more nuanced but also potentially duplicating questions in SF36.
5. Consensus that collecting both generic and AF-specific QoL would be useful in clinical practice to help clinicians understand the patient’s perspective; on balance, the group favoured using EQ-5D-5L with AFEQT.

A summary of relevant quotes from patients relating to each of the three QoL tools are presented in **Table 2**.

**Table 2:** Measurement tools for QoL in AF.

| Questionnaire | Quote | Comments/context |
| --- | --- | --- |
| SF36 | “Often I felt I fell between two tick boxes”.<br>“When you actually fill it in it gives you time to think of what you felt like. It can be the mood you’re in”.<br>“My partner coaxed me into answering, and I said do you agree with what I am saying? She responded with No, I don’t agree with that”.<br>“Pigeonholing [how I feel] into the right box that most accurately describes how I feel about my AF symptoms is difficult”.<br>“I think if I’ve got AF and there’s a risk of a stroke how can I put my health down as good?”.<br>“I think there’s a lot of duplication.....are you worn out? Have you felt downhearted? Are you full of energy?”. | There was a general consensus within the group that a diagnosis of AF led to symptoms and activity limitation that varied from day to day, making it challenging to average over a four-week period.<br>Participants commented it was good to have time for reflection when completing the questionnaire. It provided them with the opportunity to see both positive and negative aspects of their general health and wellbeing in the context of their AF diagnosis.<br>The perceptions of the carer/partner were not necessarily in agreement with the patient when completing the questionnaire.<br>The duplication of questions was raised repeatedly, leading to concern about validity from the patients. |
| EQ-5D-5L | “With this questionnaire I want to write on everything: ‘Yes but this has nothing to do with my AF’. But how is this going to be interpreted, what use will it be and am I screwing this up by filling this in this way”.<br>“I have no problem filling it in but separating AF symptoms from other health problems is difficult”.<br>“You can come up with different answers within minutes, let alone today, tomorrow and the next week”.<br>“I don’t know which [symptoms] belong to AF and I don’t know which ones belong to the other problems I have”. | Consensus was reached that this questionnaire would be useful alongside AFEQT in clinical practice to provide more accurate feedback on general health.<br>The brevity of the questionnaire was favoured, but some patients felt it difficult to sum up their well-being for that particular day.<br>As with the other questionnaires, the issue of separating co-morbidities and the general aging process was problematic.<br>Transient symptoms created difficulty when completing the questionnaire. |
| AFEQT | “The questions asked provided reassurance that my symptoms are not just about the aging process”.<br>“It makes you realise it’s not you, it’s just part of you that the AF is doing all these to you”.<br>“Filling in the questionnaire covering four weeks is difficult. Some mornings I can get up and feel fine then two hours later I feel yuck”.<br>“How do I answer [whether I am bothered]? Is it that I am not bothered at all because it’s there, or I’m very bothered because it worries me?”.<br>“Awkward questions for me... is it [due to] my medication or is it just generally how I am [with my AF]?”. | The group were unanimous in their agreement that this questionnaire was easier to complete than SF36, and gave the opportunity for informed choices within their answers.<br>The scale of being ‘bothered’ created difficulty with participants being unsure how to interpret their anxieties around their AF diagnosis and the resultant symptom burden.<br>AFEQT does not encompass general health, which in some patients had a bigger impact on overall QoL.<br>Confusion around symptoms being as a result of medications for AF, or other co-morbidities (e.g. breathlessness due to asthma). |

### (Domain 3) Importance of QoL in the management of patients with AF

The major discussion points raised in the focus groups as to whether QoL was an important outcome that clinicians should address were:

1. Improvement in QoL was the most important consideration for this patient group, ahead of mortality or the need for hospital visits.
2. Healthcare professionals in prior consultations often prioritised issues that were important to them rather than the patient, such as deciding on stroke prevention or rhythm control treatments, or on a specific target for heart rate control.
3. A lack of medication review in patients on long-standing treatments contributed to worse QoL and emotional/physical impact from AF.
4. A focus on QoL improvement could engage patients leading to empowerment. The approach in the RATE-AF trial of providing patient education and support was unanimously well received by patients in the focus groups who expressed a sense of confidence and better ability to self-manage their AF.
5. Participation in the focus group itself was beneficial for the patients, who recommended a similar process of integrated care for patients newly receiving treatment for AF, for example after discharge from hospital.

A summary of relevant quotes from patients are presented in **Table 3**; in general these reflect a mixture of the psychosocial and physical impacts of AF, coping mechanisms, and outcomes related to these effects. Whilst this study cannot address whether attention to QoL would improve clinical outcomes, there was consensus from patients that it could improve the care pathway in patients with AF.

**Table 3:** Importance of QoL for management of AF.

| Topic | Quote | Comments/context |
| --- | --- | --- |
| Ranking of outcomes | “If we’re talking about AF [the important factor] is quality of life and to be as symptom free as we can; to live each day in a way to our maximum potential whatever age we are and whatever else we’ve got, but being able to do life to our maximum”.<br>“It would be nice for the doctors to ask what is the quality of your life, how is this affecting the quality of your life”.<br>“I want a longer life and a better quality life”.<br>“A focus [on QoL] definitely had a positive impact... I don’t get out of breath like I was six months ago”. | QoL consistently rated as the most important outcome in the focus groups.<br>Patients were less concerned about other outcomes, such as hospital admissions, which were usually considered as well-managed by healthcare professionals.<br>In contrast, specific questions from healthcare professionals about the impact of AF on QoL was rare. |
| Patient empowerment | “When you go to bed of a night and you think to yourself ‘well I had a good day, I enjoyed reading that book’, or ‘at least I cleaned the stove, I did it with a little mop’. It’s ridiculous but that’s it, you go to bed [and] tomorrow is another day. If I wake up I’ll see what else I can do”.<br>“I remember standing when I was out of breath, looking in shops and pretending. I remember a man said to me one day ‘You’re out of breath’ [and I said] ‘No, I’ve got a bad back’. I was so embarrassed to say I was out of breath... and it was to do with my heart.”<br>“When you’re feeling breathless you step back and stop. [But now] I just push it that little bit further, and a little bit further, and I felt that has really helped.” | Uncertainty around living with AF, and the variability in day-to-day symptoms increased the importance of attention on QoL.<br>Lack of knowledge and fear of the unknown had a negative impact on patients.<br>Being provided with broader education (such as safely increasing exercise to work through breathlessness) was transformative for empowering patients. |
| Treatment expectations | “I don’t expect a miracle, but I would like to think that the quality of life I will have will be better because of [medications I take]”.<br>“I’ve been on the same medication for ten years. I’ve come [on this trial] and I’ve had a change... looked at thoroughly and properly, and now I feel so much better.”<br>“I could not have carried on living the life I was living.”<br>“Having been on the trial it gives you more reassurance that you’re dealing with people who are interested in AF, and I think it makes you more reassured, gives you more confidence I think. I’m happier.”<br>“I was so tired. But [now] that seems to have gone, so that’s better quality of life, and its fantastic”.<br>“I want to be treated as a person not just as an AF [patient]; I want the whole thing treated.”<br>“It helps when you know somebody else is going through it”. | Although doctors give medications in AF for specific purposes (e.g. drugs to control heart rate control), the expectation from patients is that these will improve QoL.<br>In patients with long-standing AF, patients felt that they were left without clinical attention; a refocused appraisal of medications in the context of QoL could be helpful at regular reviews.<br>Prior consultations mostly focused on stroke prevention and oral anticoagulation.<br>Being able to discuss the impact of AF with other patients in a supported group improved confidence in self-management. |

## Discussion

This study confirmed that patients with symptomatic permanent AF have considerably worse QoL than the population, or those with longstanding illness. In focus groups, QoL and symptom management were the predominant concerns of these patients. The wide variation in symptoms experienced, both generic and AF-specific, were underscored by a substantial emotional burden on patients and their families. A reported lack of focus on QoL in prior healthcare consultations contributed to a loss of confidence and a sense of isolation that hindered medical management of their condition. Further, patients felt that the extent of their comorbidities was often neglected. To improve patient well-being, management protocols need to consider other conditions such as large-joint arthritis, and not just concentrate on anticoagulation therapies for AF.

There is increasing recognition that QoL and symptom data should be collected in routine clinical practice for patients with AF^15^; however, measurement of QoL in patients with AF is associated with a number of methodological challenges. In a systematic review of measurement properties for AF-specific QoL tools, we identified substantial validity concerns for commonly-used questionnaires.^16^ Hence many research studies have used generic QoL tools such as SF36 and EQ-5D-5L. Whilst generic questionnaires allow for comparison across different diseases, they lack attention on the types of symptoms that can impair QoL in patients with AF. In our focus groups, there was consensus amongst patients that a short generic tool such as EQ-5D-5L, in addition to an AF-specific tool such as AFEQT, provided the best balance of time taken versus information gained. Although SF36 is comprehensive, and from a technical standpoint appears to cover many of the concepts discussed by patients, there was concern around difficultly to separate emotional and physical impacts from AF, the need for questions to be explained by clinical staff, and duplication in responses. An added advantage of EQ-5D-5L is the use within health economics such as quality-adjusted life-year (QALY) analyses. Whichever QoL tool is chosen, our patient groups were clear that they could see a potential benefit in rolling-out these questionnaires into routine clinical practice. Not only would this give patients a framework for their consultations with healthcare professionals, but would also allow staff to monitor changes in QoL in response to treatment.

Consideration of patient-reported outcomes is now part of international practice guidelines on AF management; clinicians are asked to assess and re-assess QoL, with symptomatic improvement a major treatment objective in AF patients.^9^ Aside from oral anticoagulation for stroke prevention, nearly all other treatments (including rate and rhythm control) are based on evaluation of symptoms. More research on assessment of QoL in clinical practice is clearly warranted to inform clinical decision making, and to tailor care to the needs of individual patients. Electronic capture of these data in routine practice could allow for real time monitoring, flexible and responsive scheduling of hospital appointments, early detection of problems, and prompt intervention to prevent AF-related adverse outcomes.^17^ Attention on comorbidities is also an essential component for optimal management of AF. In the RACE III trial, 245 patients with early persistent AF and mild-to-moderate heart failure were randomised to either targeted therapy of underlying conditions, or normal care including rhythm control.^18^ Comorbidities were better treated in the intervention group and recurrence of AF was lower (75% of patients were in sinus rhythm at 1 year, compared to 63% in the conventional management group; p = 0.042).

Similarly, integrated treatment programmes that improve patient and healthcare staff education, along with nurse-led and lifestyle interventions, have demonstrated improvements in clinical outcomes. In a randomised trial of 712 patients with AF, integrated care led to better adherence to guidelines.^19^ The composite of cardiovascular hospitalisation and cardiovascular death was significantly lower in the integrated care arm, 14.3% compared with 20.8% with usual care (hazard ratio 0.65; 95% CI 0.45–0.93; p = 0.017). Bringing together patients and their healthcare professionals in order to make shared decisions is a key element in patient empowerment^9^, especially in this digital era^20^, and has the potential to address the persistently poor outcomes seen in older patients with AF.^1^ Randomised trials to clarify the impact of AF education for healthcare professionals are ongoing.^21^ As demonstrated in a mixed-methods study of 101 patients and 15 clinicians, we can appreciate that a lack of appropriate education also hinders the ability of patients to self-manage their AF.^22^ The importance of a good knowledge-base has been studied previously, mostly in the context of anticoagulation for stroke prevention.^23^ Our impression is that education on other aspects of care, for example heart rate control, is often neglected in clinical practice. This is particularly the case for patients with permanent AF, who often receive no other therapy and are therefore left without the support that patients with paroxysmal AF typically receive when considered for rhythm control therapy.

An unexpected outcome from the focus groups was the benefit the groups themselves had for individual patients. Having a safe space to talk to other patients led to amelioration of the sense of isolation and lack of knowledge. As with other chronic diseases, many of the impacts of AF are psychosocial in nature and so better adaptation can be a powerful tool (a clear, cross-cutting theme across the domains we investigated). More widespread use of patient support groups within secondary care could have profound benefit for patients, and lead to reductions in healthcare utilisation. For example, in hospitalised patients accredited information could be provided to all patients before, during and after discharge, along with details of a national or local patient organisation for information and emotional support. Local patient support groups are run by charities such as the Arrhythmia Alliance and AF Association (https://www.heartrhythmalliance.org/), who also moderate online forums with thousands of members, organise Patient Educational Days and provide dedicated helplines. Close partnership with these organisations has the potential to support patients and their carers, and further enhance health-related QoL in those with AF.^24^

### Strengths and limitations

All included patients had permanent AF with symptom-related impairment of daily life at baseline (NYHA class II or above); hence the results presented reflect this patient group. The focus groups took place after control of heart rate and symptoms to better represent the broader community with managed permanent AF. We are limited by the number of patients involved and the need to provide depth over breadth of concepts. Greater numbers within each focus group would have limited discussion, and hence each meeting was restricted to 10 patients. We divided the sessions into two segments to avoid overloading the participants. Meetings were of sufficient duration to achieve saturation within each domain, and were undertaken within a few weeks of each other to minimise any change over time. All meetings were scheduled in a comfortable and secluded area of the hospital research unit, well known to the patients to limit anxiety. Our analytical methodology used the broad concept of thematic analysis, with a pragmatic approach to reflect that patient and public representatives were leading the focus groups, coding transcripts and providing data analysis. This provided a unique insight into patient views at many levels. Although the included patients were typical of those with permanent AF and symptom-related impairment of daily life treated in routine clinical practice, we cannot exclude distinct themes arising within patients that agreed to contribute to the focus groups. Those that participated may place a different value on the importance and subsequent assessment of QoL. However, the main discussion points raised in this research were similar in both of the randomised treatment arms, which underwent separate focus group meetings. Our findings would not necessarily apply to AF patients of other ethnic backgrounds, as there are known differences in the presentation and outcomes of AF amongst different racial groups.^25^

## Conclusion

Assessing and measuring improvement in quality of life and symptoms is fundamental to better management of patients with permanent AF, who suffer from substantial reduction in their physical wellbeing. The impact of comorbidities is poorly appreciated, with considerable variability in QoL requiring both generic and AF-specific assessment. Broader education is required beyond what is typically given around the prevention of stroke. More widespread use of questionnaires and peer support could have benefits in patients with AF seen in routine clinical practice, enabling care to be tailored to their individual needs.

## Data Availability

As per the funders position on sharing of research data, all requests for data should be directed to the Sponsor via the corresponding author. Data access requests from a third party will be managed following the relevant policies and practices of the institution hosting the research, which are expected to be transparent, robust, fair and demonstrate that appropriate mechanisms are in place to provide assurances as to the integrity of the research data. Release of data will be subject to a data use agreement between the institution and the third party requesting the data, which should detail the agreed use and appropriate management of the research data to be shared and include a requirement for recognition of the contribution of the researchers who generated the data and the original funder (UK National Institute for Health Research; policies available at www.nihr.ac.uk).

## Acknowledgments

We would like to thank other members of the RATE-AF team, in addition to the independent members of the trial oversight committees and Jonathan Mathers (Qualitative and Mixed Methods Applied Research, University of Birmingham). We are indebted to the patients and their families who dedicated their time to take part in this NHS research.

## Authors’ contributions

The Patient and Public Involvement team led the focus groups and all data extraction and thematic analysis (JJ and MS). The concept of the focus groups was originally developed by DK (Chief Investigator for the RATE-AF trial) and MC (expert in patient-reported outcomes). The manuscript was drafted by JJ, MS, KB and DK, and all authors contributed to revision and editing for intellectual content.

